# Resilient SARS-CoV-2 diagnostics workflows including viral heat inactivation

**DOI:** 10.1101/2020.04.22.20074351

**Authors:** Maria Jose Lista, Pedro M. Matos, Thomas J. A. Maguire, Kate Poulton, Elena Ortiz-Zapater, Robert Page, Helin Sertkaya, Ana M. Ortega-Prieto, Aoife M. O’Byrne, Clement Bouton, Ruth E Dickenson, Mattia Ficarelli, Jose M. Jimenez-Guardeño, Mark Howard, Gilberto Betancor, Rui Pedro Galao, Suzanne Pickering, Adrian W Signell, Harry Wilson, Penelope Cliff, Mark Tan Kia Ik, Amita Patel, Eithne MacMahon, Emma Cunningham, Katie Doores, Monica Agromayor, Juan Martin-Serrano, Esperanza Perucha, Hannah E. Mischo, Manu Shankar-Hari, Rahul Batra, Jonathan Edgeworth, Mark Zuckerman, Michael H. Malim, Stuart Neil, Rocio Teresa Martinez-Nunez

**Author notes:** All these authors contributed equally to the completion of this work (Maria Jose Lista, Pedro M. Matos, Thomas J. A. Maguire, Kate Poulton).

## Abstract

There is a worldwide need for reagents to perform SARS-CoV-2 detection. Some laboratories have implemented kit-free protocols, but many others do not have the capacity to develop these and/or perform manual processing. We provide multiple workflows for SARS-CoV-2 nucleic acid detection in clinical samples by comparing several commercially available RNA extraction methods: QIAamp Viral RNA Mini Kit (QIAgen), RNAdvance Blood/Viral (Beckman) and Mag-Bind Viral DNA/RNA 96 Kit (Omega Bio-tek). We also compared One-step RT-qPCR reagents: TaqMan Fast Virus 1-Step Master Mix (FastVirus, ThermoFisher Scientific), qPCRBIO Probe 1-Step Go Lo-ROX (PCR Biosystems) and Luna^®^ Universal Probe One-Step RT-qPCR Kit (Luna, NEB). We used primer-probes that detect viral N (EUA CDC) and RdRP (PHE guidelines). All RNA extraction methods provided similar results. FastVirus and Luna proved most sensitive. N detection was more reliable than that of RdRP, particularly in samples with low viral titres. Importantly, we demonstrate that treatment of nasopharyngeal swabs with 70 degrees for 10 or 30 min, or 90 degrees for 10 or 30 min (both original variant and B 1.1.7) inactivates SARS-CoV-2 employing plaque assays, and that it has minimal impact on the sensitivity of the qPCR in clinical samples. These findings make SARS-CoV-2 testing portable to settings that do not have CL-3 facilities. In summary, we provide several testing pipelines that can be easily implemented in other laboratories and have made all our protocols and SOPs freely available at https://osf.io/uebvj/.

## Introduction

“Test, test, test” – this was the message from the World Health Organization’s Head Tedros Adhanom Ghebreyesus on the 16^th^ of March 2020. This message is still current, more than a year after the pandemic was declared. To fight the exponential spread of SARS-CoV-2, measures of social distancing have been imposed in many countries worldwide, while others are now in a phase of de-escalation. Social distancing and lockdown measures have resulted in stagnant or dropping numbers of new infections-however, appearance of outbreaks has proven inevitable in places where measures have been relaxed. Vaccination has decreased the spread in certain countries; however, these are few nations and the spread of new variants makes testing as imperative as before. Test, Trace and Isolate have been essential to halt SARS-CoV-2 infection. Non-PCR tests such as lateral flow tests have proven useful particularly in the case of symptomatic testing^1^; however, large scale PCR-based testing is essential to contain and prevent outbreaks due to its high sensitivity. This is particularly relevant in asymptomatic individuals and should also be central in implementing an ‘exit strategy’ plan.

In order to increase testing capacity, many countries rely on centralised efforts to build large diagnostic centres. However, the involvement of smaller academic or commercial laboratories has proven helpful and necessary too ^2-5^. These decenatralised laboratories can repurpose existing equipment and technical expertise for SARS-CoV-2 testing, for example by comparing methods of extraction vs extraction-free methods or samples treated with heat^6-11^, combining heat with proteinase K treatments to improve sensitivity^12^ or establishing sensitivity of primer-probe pairs^13,14^. The UK government document “Guidance for organisations to seek supporting the COVID-19 testing programme” published on the 9^th^ of April 2020, by the Department of Health and Social Care, clearly welcomes academic institutions to increase testing capacities within the UK referred here as NHS-helper labs. However, due to global high demand of the kits and reagents used in the WHO (World Health Organization), CDC (Centre for Disease Control, US), ECDC (European Centre for Disease Prevention and Control) and PHE (Public Health England) ratified testing strategies, the NHS-helper labs are encouraged to use alternative strategies that will not interfere with the reagent demand of larger testing facilities. Moreover, helper laboratories can provide their research expertise and experimental validation of other kits enabling clinical labs to benefit from their results. We set out to perform this task.

Here we describe different strategies for SARS-CoV-2 PCR-based detection by employing reagents that are not currently used in the NHS setting. We also performed heat inactivation employing dry bead baths of SARS-CoV-2, for both the original strain and the more recent B 1.1.7 strain and present data supporting good limit of detection (LoD) for both strains upon heat treatment. Within the UK, the NHS agrees on the use of alternative RNA isolation and qPCR protocols, providing these have been internally validated and discussed with the local NHS partner. To increase visibility of these alternative strategies, we have created a webpage under the Open Science Framework platform (https://osf.io/uebvj/) that we hope will stimulate exchange between smaller laboratory facilities, increase confidence in tested alternative routes of RNA isolation and viral RNA detection and thereby expedite the establishment of smaller academic testing centres.

## Results

Our pipelines are adaptable for both manual and automatic handling; we also employ heat inactivation of virus within the swabs for easier processing. We compare three RNA extraction methods, one column-based and two magnetic beads-based that are currently or have the potential to be automatized. As a benchmark, we use the QIAamp Viral RNA Mini Kit (QIAGEN) as their proprietary buffer AVL inactivates SARS-CoV-2 according to CDC guidelines. We also validate three different one-step RT-qPCR kits. We use the CDC recommended N1 and N2 primer-probe sets^15^ and compare these against the PHE recommended RdRP_SARSr primers^16^. We did not test efficiency of the reverse transcription (RT) step, as we had no access to *in vitro* transcribed RNA. For these validations we received clinical swab material from St Thomas’ Hospital and King’s College Hospital (London, UK) and compared our results with their diagnostics clinical pipelines. Detailed step-to-step standard operating procedures (SOPs) can be found at https://osf.io/uebvj/.

We have created a flowchart of the different processing steps and combinations in our pipeline (Figure 1), which we subsequently explain in more detail.

**Figure 1.**
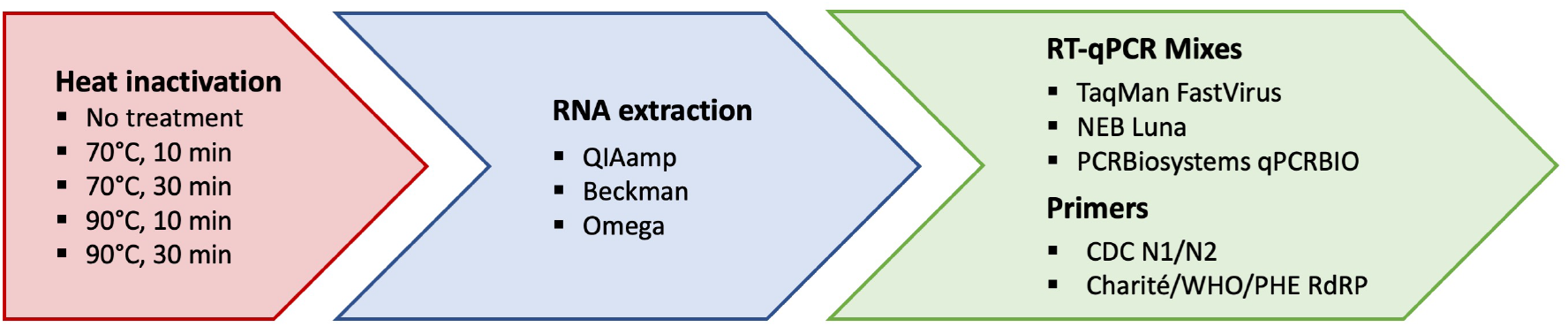
Representation of our workflow. We employed heat inactivation vs non heat inactivation ^24^; compared three different RNA extraction kits (blue) followed by three RT-qPCR mixes and three sets of primers (green).

To test the efficiency and detection range of the CDC-recommended N1 and N2 primer-probes, we amplified serial dilutions of plasmids encoding the N SARS-CoV-2 gene (positive controls provided by Integrated DNA Technologies, IDT) using the TaqMan™ Fast Virus 1-Step Master Mix (Figure 2A), FastVirus hereafter. Good linearity could be achieved up to 10 copies of DNA molecules. Using the N1 and N2 primer-probes, we compared the efficiency of RNA recovery between the column-based QIAamp Viral RNA Mini Kit (QIAGEN, QIAmp herein) endorsed by the CDC, and two magnetic bead extraction kits: the RNAdvance Blood (now RNAdvance Viral) (Beckman hereinafter) and Mag-Bind Viral DNA/RNA 96 Kit (Omega Bio-tek, Omega herein), starting from the same material (140 μL). RNA isolation from four different coronavirus positive samples (CPS) with all three kits rendered comparable cycle thresholds (Cts) when amplified with the primer-probes N1 and N2. This was the case for two different RT-qPCR Master Mixes, FastVirus (Figure 2B) or Luna^®^ Universal Probe One-Step RT-qPCR Kit (NEB, Luna hereafter) (Figure 2C).

**Figure 2.**
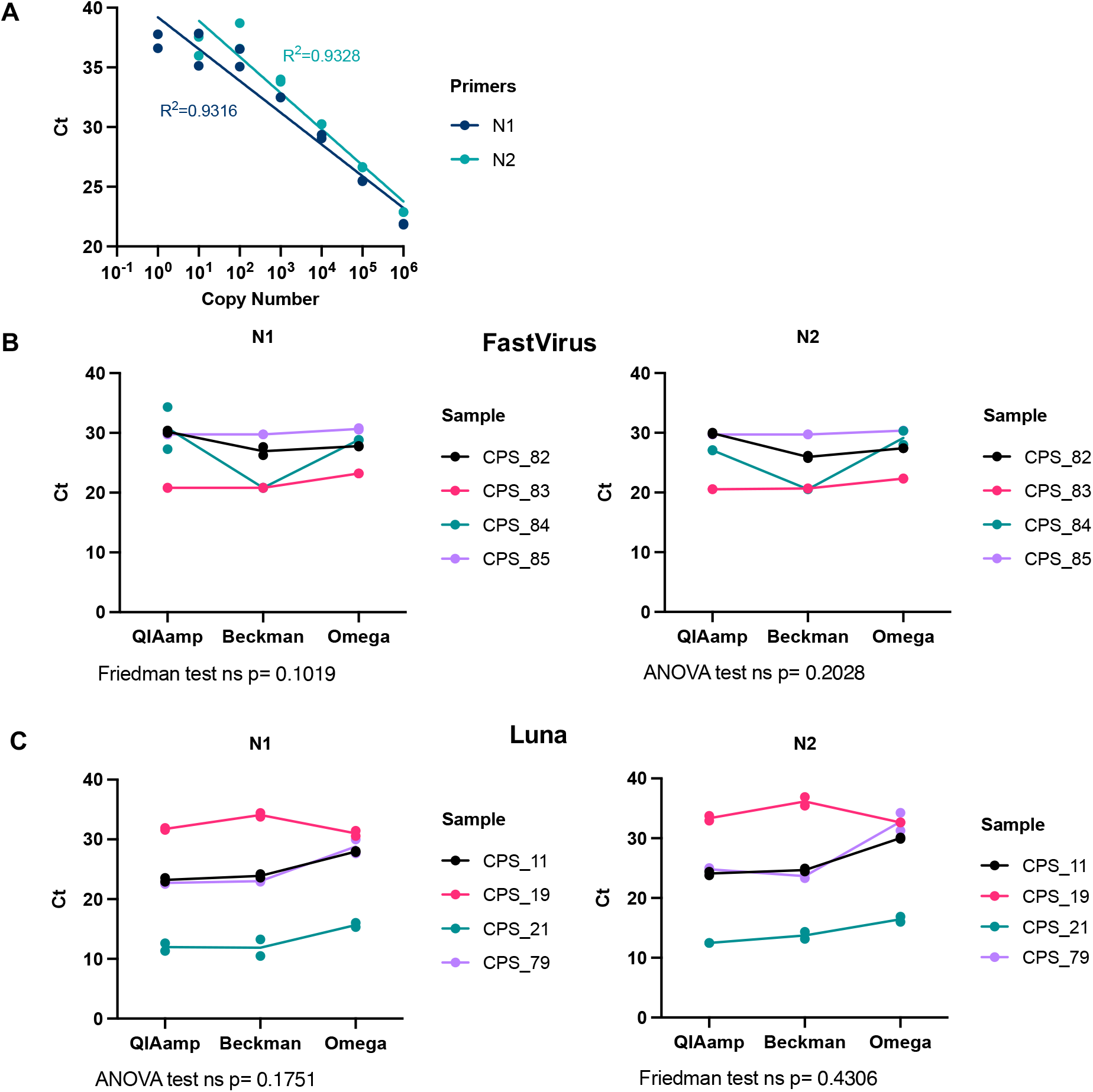
Comparison between different RNA extraction methods. **(A)** Dilution curve of the positive control provided by IDT (plasmid containing SARS-CoV-2 N gene) using N1 or N2 primer-probe sets with the Taqman FastVirus mix. A semi-log regression was used to calculate the coefficient of determination (R^2^). **(B, C)** A set of four swab samples were used for RNA extraction with the indicated kit. RT-qPCR was run with N1 and N2 primer-probe sets employing the FastVirus **(B)** or the Luna Master mixes **(C)**. Shapiro-Wilk test was used for normality assessment prior to analysis employing ANOVA (parametric data) or Friedman test (non-parametric data). These samples were previously classified as positive (CPS) by the diagnostics lab, the number indicates different donors. Dots represent each individual RT-qPCR technical duplicate, line connects average of replicates and statistics were performed in average duplicates.

We also compared different one-step RT-qPCR kits to amplify swab-material purified using the QIAamp viral RNA mini kit which we considered our ‘benchmark’ given CDC-guidelines on buffer AVL inactivating SARS-CoV-2. Supplemental Figure 1 shows RNA from ten different positive donors amplified with Luna, FastVirus and qPCRBIO Probe 1-Step Go Lo-ROX (PCR Biosystems, qPCRBio). All Master Mixes detected comparable amounts of RNA using primer-probes against N1 primer-probe, with the exception of donor CPS_101 which had borderline Ct values of 38 in both FastVirus and Luna and was undetectable using qPCRBio Master Mix (ANOVA p=0.1278). Tukey’s multiple comparisons showed Luna performing better than FastVirus (p adjusted=0.007) with no samples missed by FastVirus. Thus, three different RNA extraction kits, and three different one-step RT-qPCR kits achieve detection of viral RNA within swab material.

As a diagnostic assay, it is paramount to be able to detect very low viral loads in swab samples. To determine the efficiency of the RT-qPCR we serially diluted the RNA from a confirmed positive swab isolated with each one of the three different kits used in this study. All dilutions were assessed with the N1 and N2 primer-probe amplification employing the Luna Master Mix. Figure 3A shows that the RT-qPCR reaction remains linear over a 10^5^-fold RNA dilution range. To ensure that low viral presence can be reliably detected with each one of these methods, we prepared serial dilutions of swab material from three different CPS donors in Hank’s Balanced Salt Solution (HBSS) + 1% bovine serum albumin (BSA) since viral transport medium contains these only with the addition of amphotericin and gentamicin. Viral RNA was isolated from these diluted swabs with the three RNA isolation kits from Figure 2. Figure 3B shows that all three kits recover viral RNA over a wide range of concentrations, with the N gene being reliably amplified with the N1 and N2 primer-probe sets and the Luna Master Mix. CPS21 10^−1^ dilution was excluded from the r calculations. The N2 primer-probe set in the donor CPS79 extracted with Omega showed poor linearity, possibly related to initial variation in the non-diluted sample.

**Figure 3.**
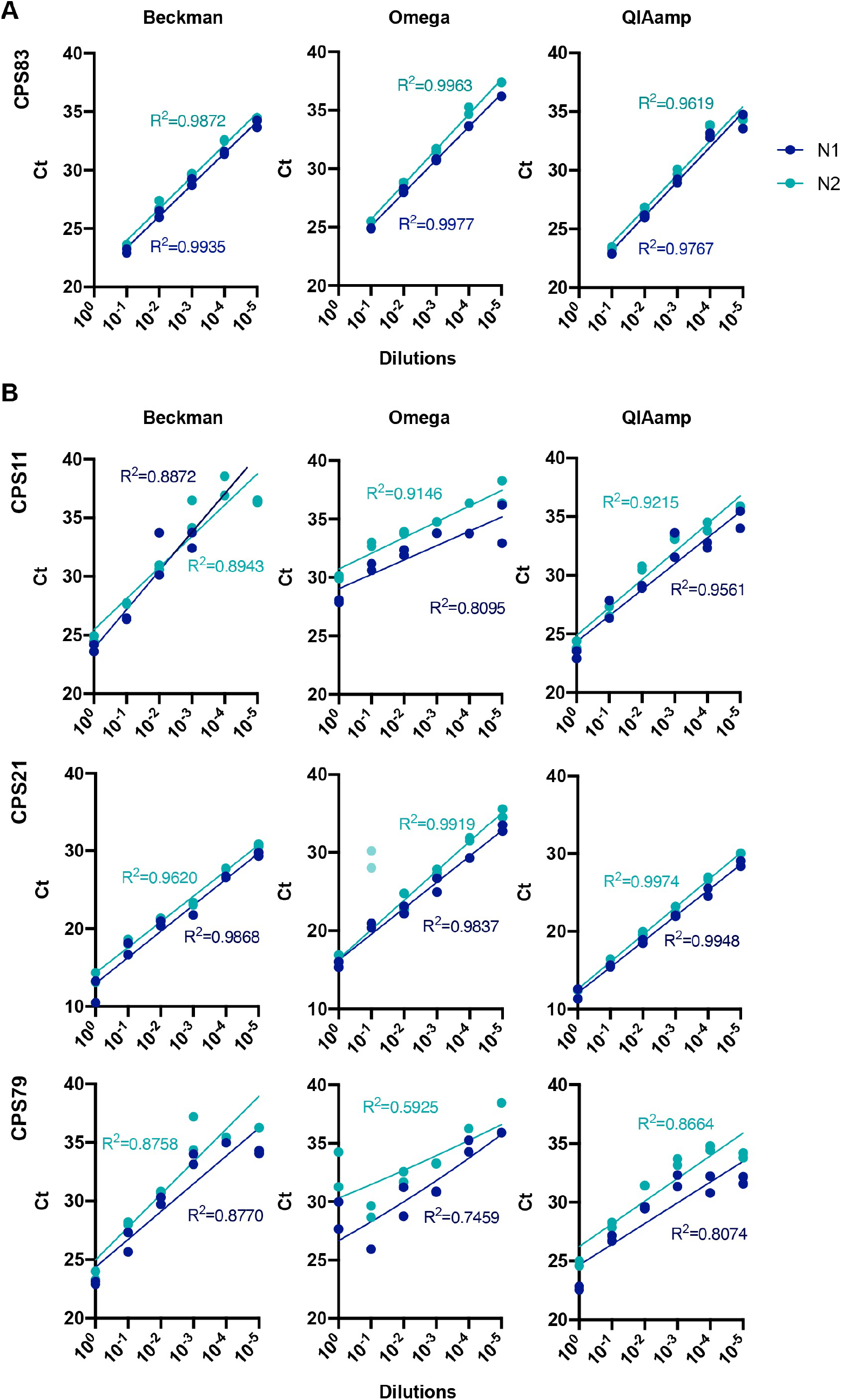
Sensitivity of qPCR detection by serial dilutions of extracted RNA or swab samples. **(A)** RNA from sample CPS83 was serially diluted and extracted with three different methods. RT-qPCR was run with N1 and N2 primer-probe sets with Luna Master Mix. **(B)** Three distinct positive swab samples (CPS) were serially diluted followed by RNA extraction by the indicated method. RT-qPCR was run with N1 and N2 primer-probe sets with Luna Master Mix. Dots represent each individual technical duplicate. A semi-log regression was used to calculate the coefficient of determination (R^2^).

One major limitation for many academic and commercial laboratory settings is the lack of available CL-3 laboratory space and/or Class I MSCs required to handle/open the potentially infectious swabs. Moreover, samples with high viral load pose a risk of infection for the handler. Most recommended viral inactivation protocols use a combination of guanidinium thiocyanate (GTC) and Triton X-100. GTC became scarce due to its wide use, it is quite toxic and not compatible with some kits, and the use of chemical inactivation protocols after sample collection inherently requires opening of the swab sample, with the consequent risk of exposure for lab staff. Heat treatment of viral particles has been shown effective in inactivating SARS-CoV-2 with 70°C 5min treatment rendering viral infectivity undetectable employing Vero-6 cells (limit of detection of TCID50 assay was 100 TCID50/mL)^17-19^. Other heat treatment protocols have also been demonstrated, with variable effects on PCR sensitivity^20^.

We firstly assessed different temperature and time conditions for heat treatment of both SARS-CoV-2 original and B 1.1.7 strains. The novel strain of SARS-CoV-2 B.1.1.7 was originally described in December 2020 in the UK, firstly detected in samples as early as 20^th^ September 2020^21^. Since then, it has spread to many other countries where it is now the predominant strain. We thus evaluated the effect of heat in the detection of B.1.1.7. To that end we performed serial dilutions of cultured virus in viral transport medium, extracted employing Beckman and assessing the presence of N using the N1 and N2 primer probe combinations in the resulting RNA. Figure 4 shows that heat at either 70°C (4A) or 90°C (4B) for 30 minutes did not alter the detection of viral RNA (red vs blue). Figure 4C and Supplemental Figure 2A show that treatment of B.1.1.7 with 70°C or 90°C for 10 and 30 mins fully inactivates this strain of SARS-CoV-2 virus. Supplemental Figure 2B shows complete inactivation of the original strain at all temperatures and times. Importantly, we observed incomplete inactivation when temperatures between 60 and 70 degrees were applied for 30 min (Supplemental Figure 3), making imperative the use of well-calibrated thermometers, or use of higher temperatures/longer time periods.

**Figure 4.**
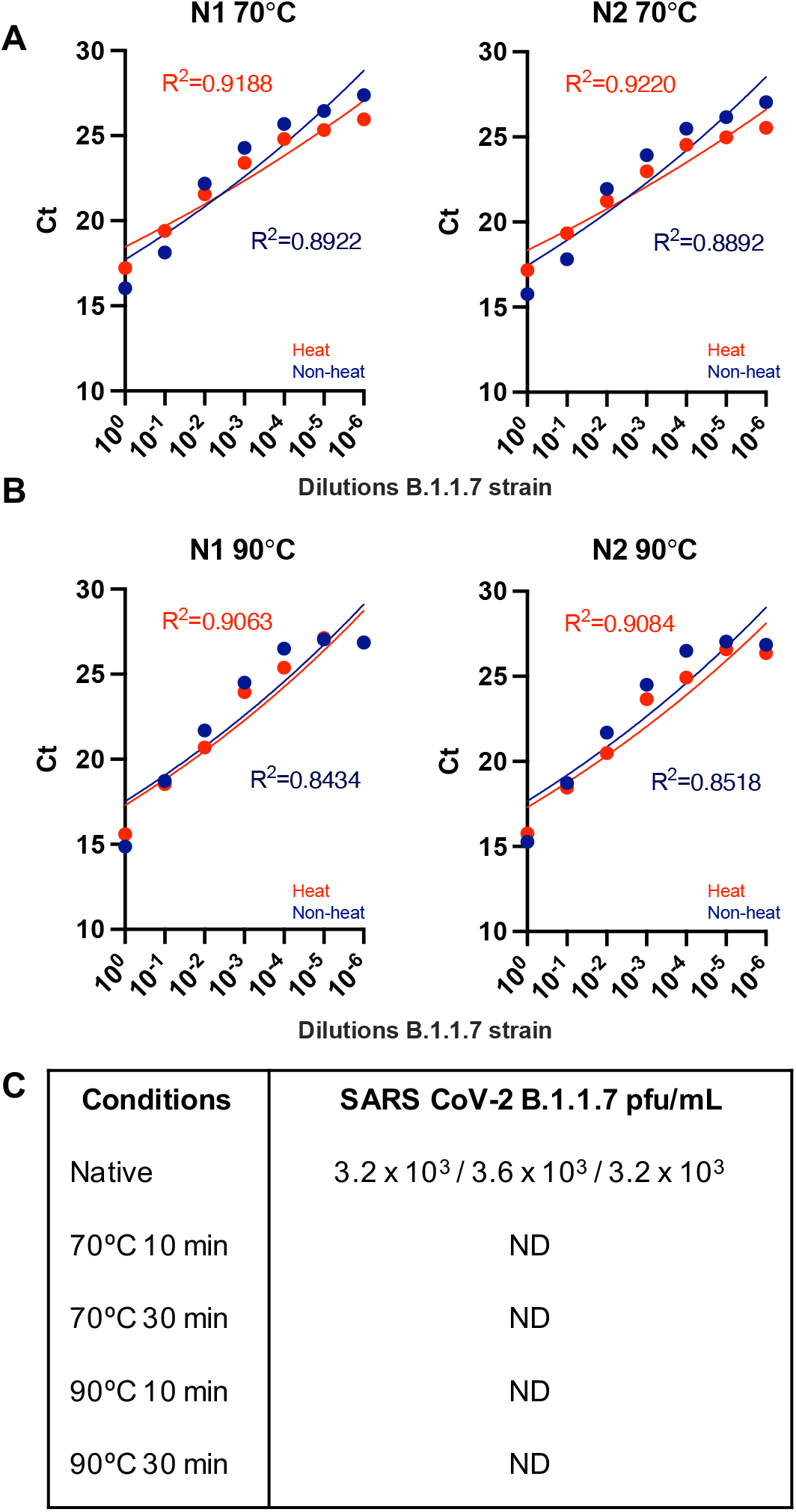
Sensitivity of qPCR detection by serial dilutions of viral stocks of the new B.1.1.7 strain. B.1.1.7 SARS-CoV-2 viral stocks were serially diluted in viral transport medium, extracted employing Beckman and assessed employing N1 and N2 primer-probe sets using the FastVirus Master Mix. Samples were heat treated for 30 minutes with either 70°C **(A)** or 90°C **(B)**. Dots represent the mean of the qPCR technical duplicates. A semi-log regression was used to calculate the coefficient of determination (R^2^). **(C)**. Results of plaque assays for heat treatment of cultured SARS-CoV-2 B.1.1.7 variant.

We then assessed if heat treatment of nasopharyngeal swab-material constitutes a method of treating samples within their original unopened collection tubes without compromising RT-qPCR results. Data shown were obtained employing QIAamp RNA extraction. We first assessed heating sample aliquots at 70°C for 30min with different samples and performed RT-qPCR with N1 primer-probes and Luna Master Mix. As Figure 5A shows, we did not observe any change in Ct values upon heat treatment of the sample (t-test p=0.1946).

**Figure 5.**
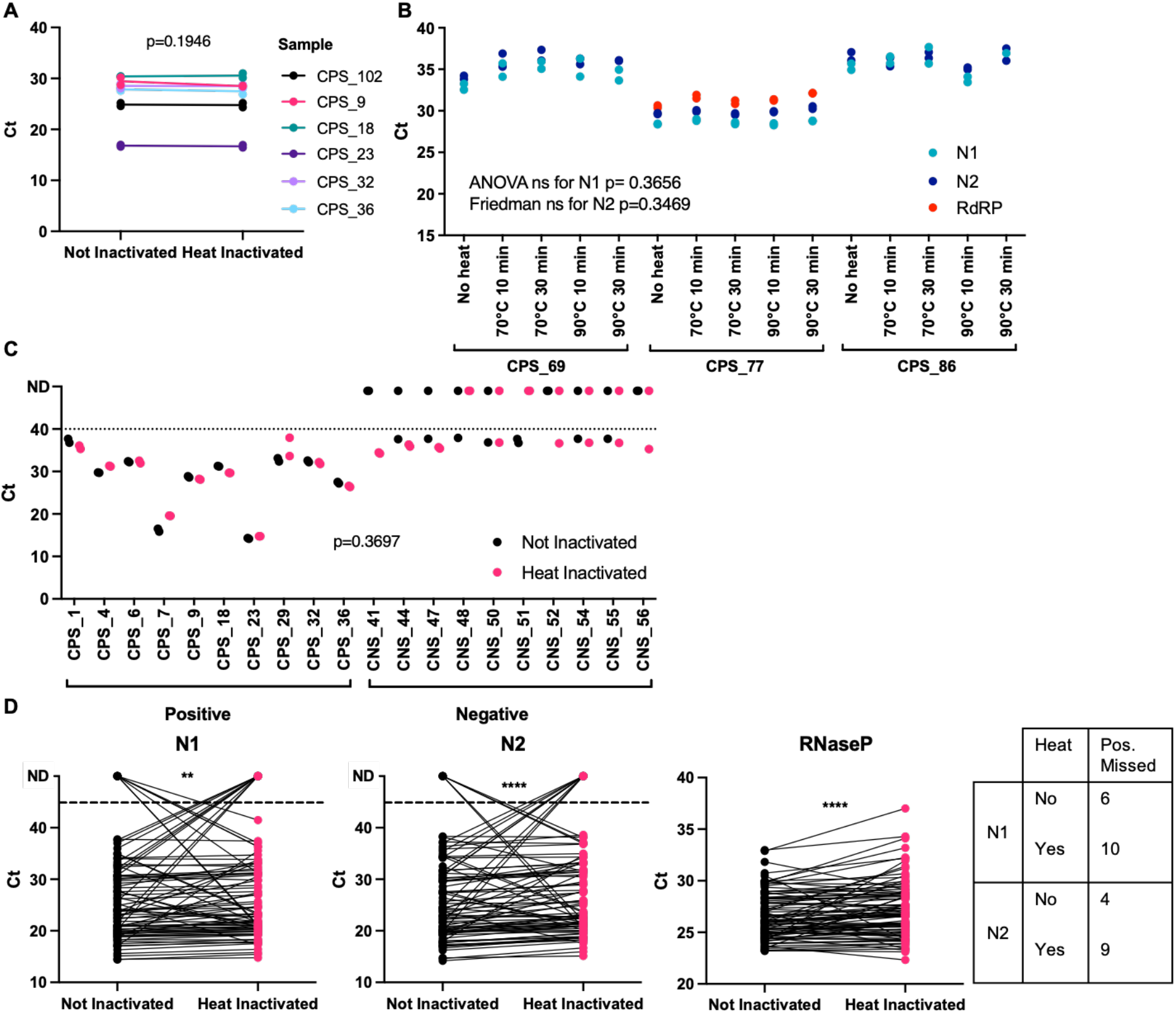
Primer comparisons and heat inactivation of nasopharyngeal swab samples. **(A)** A different set of six positive samples were used to compare directly non treated with heat treated at 70°C for 30 min. RNA extraction was done with QIAamp and RT-qPCR with N1 primers and NEB Luna mix. **(B)** Three additional positive samples were subjected to different temperatures and incubation times as indicated, with RNA extracted by QIAamp. All three primer-probe sets from panel A were used, together with Taqman FastVirus master mix. **(C)** 20 additional samples, 10 from positive donors (CPS) and 10 from negative donors (CNS) were used to compare directly non treated with heat treated at 70°C for 30 min. RNA extraction was done with QIAamp and RT-qPCR with N1 primers and Luna Master Mix. For panels A-C, dots represent each individual technical duplicate, line connects the average of replicates. **(D)** 93 additional samples were non treated or heat treated at 90°C for 10 min. RNA extraction was done with Beckman and RT-qPCR with N1, N2 and RNAseP primer-probes and FastVirus Master Mix. Samples were run in singlets.

We then set out to test a wider range of heat-inactivation conditions on six confirmed positive samples using two extraction methods. We treated aliquots of the same sample with no heat, 70°C for 10min, 70°C for 30min, 90°C for 10min or 90°C for 30min and extracted RNA employing the QIAamp kit. We employed a dry metallic bead bath to heat the sample tubes, since water baths are not allowed in Cat-3 laboratories due to the risk of spillage. Our results show that none of the heat conditions altered the Ct values (ANOVA for N1 p= 0.3656 and Friedman test for N2 p=0.3469, Figure 5B). Both the N1 and N2 primer-probe sets gave reliable and near-identical amplification of viral RNA; however, we noticed that the RdRP primer-probe set failed to amplify viral samples with high Ct values. These results of high Ct values for RdRP were also observed when we used the MagMax kit (ThermoFisher Scientific) as an extra RNA extraction method (Supplemental Figure 4). Comparison of the different primer-probe combination for RdRP rendered similar results (Supplemental Figure 5) and as previously shown^13^. This important finding should be considered when laboratories decide to rely only on one primer-probe set.

To confirm the reproducibility of our results, we employed another distinct set of samples, assessing both positive and negative samples. We aliquoted swab material, warmed it at 70°C for 30min, extracted their RNA using QIAmp and performed RT-qPCR using Luna Master Mix using primer-probe N1. Ct values did not change upon heat inactivation as observed previously (t-test p=0.3697). However, we determined multiple instances where swabs previously identified as negative amplified in one of the RT-qPCR duplicates at high Ct values of around 38 (Figure 5C). All our water controls (no template and water template) yielded no amplification. RNAse P controls are in Supplemental Figure 6.

To further test a higher temperature we employed 90°C for 10min in 93 samples, extracted their RNA using Beckman and performed RT-qPCR using FastVirus Master Mix using primer-probes for N1, N2 and RNAseP. Figure 5D shows the results for those samples where we detected positive amplification in either of the treatments (88 for N1 and 90 for N2). More samples were lost upon heating, although statistical analysis of Ct ranges remained similar between heat vs non-heat (Supplemental Table 1). N2 primer-probe appeared more heat-resistant, as 90 samples were detected vs 88 with N1 primer-probes. Interestingly, we observed no clear trend for samples with high Ct values lost upon heat treatment (Supplemental Figure 7).

## Discussion

Our work shows different workflows for nasopharyngeal swab processing for SARAS-CoV-2 detection employing different combinations of inactivation, extraction and amplification. We present data employing our in-house validation of two viral RNA purification kits (Beckman and Omega) as alternatives to the QIAamp viral RNA mini kit. We have also tested three alternative, commercially available one step RT-qPCR kits (FastVirus, Luna and PCRBio) and assessed different recommended primer-probe sets (N1, N2, RdRP) which can currently detect all circulating SARS-CoV-2 variants to date.

With regards to extraction methods, our data in Figure 2 suggest that Beckman performs better as seen for sample CPS_84. This is also supported by data in Figure 3 where CPS_21 and CPS_79 samples showed no good linearity when diluted and extracted with Omega. When analysed together, we did not observed any statistically significant difference between Luna, FastVirus and PCRBio, although one sample was not detected employing PCRBio and Luna appeared to perform better than FastVirus (Supplemental Figure 1). The difference in Ct values between Luna and FastVirus was very small and did not alter overall sensitivity (mean Ct difference 0.8335).

We find that N1 and N2 primer-probe are more sensitive than RdRP (Figure 5B), similar to others^13^. This may likely be due to higher amount of sub-genomic RNAs encoding for N^22^. Our data show that heat inactivates the original and B 1.1.7 strains at 70°C or 90°C for 10 and 30 mins (Figure 4 and Supplemental Figure 2). Importantly, we observed that high viral concentrations using inadequate temperatures between 60 and 70°C are not fully inactivated (Supplemental Figure 3). We observed this in early experiments where were relied on a heat block thermometer, which was proven inaccurate and set at a temperature of ∼62-63°C instead. This highlights the need to accurately measure temperatures when performing inactivation of clinical samples – or the use of temperatures above 70°C. Importantly, temperatures of the swabs must be considered when performing heat inactivation, since swabs kept in the fridge will take longer to reach a certain temperature vs swabs kept at room temperature. Our data also highlight that adequate titrations employing high viral loads of SARS-CoV-2 are required to establish if full inactivation has been achieved. 70°C during 30 min appeared to have no effect on sensitivity (Figure 5C) while 90°C during 10 min appeared to decrease sensitivity (Figure 5D). Our data supports heating at temperatures below 90°C, method that may be used to reduce the need for CL-3 laboratory and to speed up sample processing given the chemical inactivation methods are labour-intensive increasing the risk of exposure to the lab staff. Some laboratories have implemented dry heat in ovens^4^ to inactivate samples; we propose the use of dry heat with beads as it allows for both high and low throughputs and is safe against possible sample leaks (beads can be disinfected at the moment of leakage). Our pipeline can therefore be implemented in places that only have CL-2 facilities to detect SARS-CoV-2. Our results differ from those observed by others with regards to samples with low viral loads being lost upon heating^20^ since we did not observe a clear pattern of samples with low viral loads being undetected upon heat and interestingly some samples were only detected upon heat treatment (Supplemental Figure 7). Our higher temperatures employed may possibly denature RNAses and/or facilitate viral RNA denaturation while preserving enough integrity for detection with the N1 and N2 primer-probe sets. Regardless, we advise the use of 70°C over 90°C when possible.

Together these data highlight the need for performing cross-validations of RNA extraction kits and primer-probe pairs prior to implementing in diagnostics, with an emphasis on the need of using clinical samples (rather than diluted RNA or plasmid DNA) to establish ‘real-world’ data that better relate with clinical samples. Swab material and inherent inhibitors will perform variably with different workflows and we thus highlight the need to assess their performance – a task in which diagnostics laboratories can leverage on collaborations with academic institutions to speed up the establishment of new protocols.

Based on the above, and understanding that including RT-qPCR duplicates may decrease the number of samples a diagnostic laboratory can process (particularly if employing 96 well plates), we suggest to:

- employ heat (70°C preferably to 90°C) for 10-30min. Ensure temperature is at least 70°C;
- preferably employ N1 and N2 primer-probes vs RdRP;
- test samples without RT-qPCR technical replicates to increase the testing throughput;
- run duplicates in case of borderline ≥36Ct^23^ and always check amplification curves of samples. If 1) amplification is shown reproducibly consider it a positive sample with low viral load 2) amplification unclear (one replicate positive, one negative) for these donors to be re-tested as soon as possible to confirm positive or negative detection of SARS-CoV-2 regardless of symptoms. Although we acknowledge the limitations, if possible, re-swabbing of doubtful samples is highly recommended as a first option.

## Methods

All materials with their catalogue numbers are available at https://osf.io/uebvj/.

### Heat inactivation

Swab tubes containing Viral Transport Medium (VTM) were checked for cracks to ensure no viral material had leaked. Swab tubes were then transferred to a water bath (Grant) containing dry metallic beads (Starlab) preheated to 70°C or 90°C, ensuring the entire swab tube (including lid) was covered by the beads. Samples were incubated in the following conditions: 70°C for 10 mins, 70°C for 30 mins, 90°C for 10 mins, or 90°C for 30 mins, then transferred back to Class I MSC and allowed to cool to room temperature prior to RNA extraction.

### RNA Extraction

Serial dilutions were done in Hank balanced salt solution (HBBS) and 1% Bovine Serum Albumin (BSA) to closely mimic viral transport media,

#### Qiagen QIAamp Viral RNA Mini Kit

From swab tube, 140 µl sample was transferred to 1.5 mL screw-cap microcentrifuge tube and treated with 560 µl AVE, containing carrier RNA, followed by 560 µl molecular-grade 100% ethanol (Fisher Scientific). Samples were then taken out of the Class I MSC and CL-3 lab as AVL is known to inactivate SARS-CoV-2, transferred into QiaAmp mini spin columns (Qiagen) and centrifuged according to manufacturer’s instructions. Two wash steps were performed, with 714 µl buffer AW1 and 714 µl buffer AW2 (both Qiagen). RNA was then eluted from the columns with 40 µl RNase-free water (Ambion), followed by a second 40 µl elution to maximise RNA yield and giving a final RNA sample volume of 80 µl.

#### Beckman Coulter Agencourt RNAdvance Blood Total RNA Kit

Reagents were prepared prior to RNA extraction according to manufacturer’s instructions. The protocol was conducted in a Class I MSC in a CL-3 lab. From a swab tube, 140 µl were transferred to a Zymo-Spin I-96 Plate (Zymo Research). 7 μl of Proteinase K/PK buffer and 105 µl of Lysis buffer was added to each sample, mixed and incubated at room temperature for 15 minutes. Following incubation, 143 μl of Bind1/Isopropanol was added to each sample, mixed, and the samples were left to incubate at room temperature for 5 min. The Zymo-Spin I-96 Plate was placed on ZR-96 MagStand (Zymo Research), and the magnetic beads left to form a pellet. The supernatant was removed, and the magnetic beads washed three times, first, with 280 μl of Wash buffer (Beckman Coulter), followed by two washes with 70% ethanol. Following the wash steps, RNA was eluted from the columns with 80 µl RNase-free water (Ambion).

#### Omega Bio-tek Mag-Bind^®^ Viral DNA/RNA kit

Reagents were prepared prior to RNA extraction according to manufacturer’s instructions. The protocol was conducted in a Class I MSC in a CL-3 lab. From a swab tube, 140 µl sample was transferred to a Zymo-Spin I-96 Plate (Zymo Research). 369.5 μl of Lysis mastermix was added to each sample, mixed, and incubated at room temperature for 10 minutes. Following incubation, 7 μl of Mag-Bind^®^ Particles CNR and 7 μl of Proteinase K solution was added to each sample, mixed and incubated at room temperature for 5 minutes. The Zymo-Spin I-96 Plate was placed on ZR-96 MagStand (Zymo Research), and the magnetic beads left to form a pellet. The supernatant was removed, and the magnetic beads washed three times, first, with 280 μl of VHB buffer (Omega Bio-tek), followed by two washes with 350 μl SPR Wash Buffer (Omega Bio-tek). Following the wash steps, RNA was eluted from the columns with 80 µl RNase-free water (Ambion).

### One-step RT-qPCR

#### qPCRBIO Probe 1-Step Go Lo-ROX (PCR Biosystems)

Reactions were done with 5 µL RNA, 5 µL 2x qPCRBIO Probe 1-Step Go mix, 1.2 µL forward primer RdRP_SARSr-F2 (10 µM), 1.6 µL reverse primer RdRP_SARSr-R1 (10 µM), and 0.2 µL probe RdRP_SARSr-P2 (10 µM), 2 µL of 20x RTase Go, and completed with RNase-free water to 20 µL. The samples were incubated in a QuantStudio 5 (Applied Biosystems/ThermoFisher Scientific). Reverse transcription was performed for 10 minutes at 45°C. The DNA polymerase was activated for 2 minutes at 95°C and the samples underwent 50 cycles of denaturation (5 seconds at 95°C) and annealing/extension (30 seconds at 60°C). A plate read was included at the end of each extension step. Each sample was run in duplicate.

#### TaqMan Fast Virus 1-Step Master Mix (Applied Biosystems)

Reactions were performed with 5 µL RNA, 5 µL TaqMan Fast Virus 1-Step master mix, with probes and water making the 20 µL reaction. For Charité/WHO/PHE primers, 1.2 µL forward primer RdRP_SARSr-F2 (10 µM), 1.6 µL reverse primer RdRP_SARSr-R1 (10 µM), and 0.2 µL probe RdRP_SARSr-P2 (10 µM), and 7 µL RNase-free water were used. For CDC primers (EUA kit IDT), 1.5 µL of each primer-probe premixture (N1, N2 or RNAseP) and 8.5 µL water were used. The samples were run in a QuantStudio 5 (Applied Biosystems/ThermoFisher Scientific) using the “Fast” cycling mode. Reverse transcription was performed for 5 minutes at 50°C. The reverse-transcriptase was then inactivated for 20 seconds at 95°C and the samples underwent 50 cycles of denaturation (3 seconds at 95°C) and annealing/extension (30 seconds at 60°C). A plate read was included at the end of each extension step. Each sample was run in duplicate.

#### Luna Universal Probe One-Step RT-qPCR (NEB)

Reactions were performed with 5 µL RNA, 10 µL 2x Luna Universal Probe One-Step reaction mix, 1 µL Luna WarmStart RT enzyme mix, 1.5 µL of each CDC primer-probe premixture (N1, N2 or RNAseP), and 2.5 µL RNase-free water. The samples were incubated in a QuantStudio 5 (Applied Biosystems/ThermoFisher Scientific) using the “Fast” cycling mode. Reverse transcription was performed for 10 minutes at 55°C. The samples were denatured for 1 minute at 95°C and then underwent 50 cycles of denaturation (10 seconds at 95°C) and annealing/extension (30 seconds at 60°C). A plate read was included at the end of each extension step. Each sample was run in duplicate.

Primer and probe sequences are supplied in Supplemental Material. Supplemental Tables 2 and 3 show the volume reaction and cycling conditions.

### Virus work

Original SARS-CoV-2 Strain England 2 (England 02/2020/407073) was obtained from Public Health England and 2 lineage B 1.1.7 (VOC 2 2020212/01) was kindly provided by W. Barclay (Imperial College London). The infectious virus titre was determined by plaque assay in Vero-E6 cells. Limit of detection was 40 plaque forming units (pfu)/mL.

Vero-E6 cells were kindly provided by W. Barclay (Imperial College London). Cells were maintained in complete DMEM GlutaMAX (Gibco) supplemented with 10% foetal bovine serum (FBS; Gibco), 100 U/mL penicillin and 100μg/mL streptomycin and incubated at 37°C with 5% CO2.

### Study approval

This study was approved by Guy’s and St Thomas’ NHS Trust, REC Ref 18/NW/0584

## Data Availability

All raw data files are available to anyone that requires them. Our protocols are also published at https://osf.io/uebvj/

## Acknowledgements

This work was funded by; King’s Together Rapid COVID-19 Call awards to RTM-N, MHM, KD, SN, JE and MS-H, MRC Discovery Award MC/PC/15068 to SN, KD and MHM, a Huo Family Foundation Award to MHM, KD, RTM-N, SN, JE and MS-H, MRC Programme Grant MR/S023747/1 to MHM, Wellcome Trust Investigator Award 106223/Z/14/Z to MHM, NIAID Awards U54AI150472 and R01AI076119 to MHM. A.S., M.F. and R.E.D. were supported by the MRC-KCL Doctoral Training Partnership in Biomedical Sciences (MR/N013700/1); E. O-Z was supported by MR/S009191/1 to Parsons M and Santis G; M.F. was supported by the MRC (MR/R50225X/1); H.S was supported by the BBSRC (BB/P504609/1); R.P. was supported by the by the (NIHR) Biomedical Research Centre based at Guy’s and St Thomas’ NHS Foundation Trust and King’s College London; T.J.A.M. was supported by Asthma UK at the Asthma UK Centre in Allergic Mechanisms of Asthma; P.M., S. P. and H.W. were supported by WT098049AIA to Neil S. and Swanson C.; M.J.L.B and C.B. were supported by MRC (MR/S000844/1) to Neil S. and Swanson C. This UK funded award is part of the EDCTP2 programme supported by the European Union; K.P. was funded by KHP Challenge Fund to R.T.M-N; M.H. was funded by RP_007_20190305 from Kidney research UK; H.M. was funded by the Wellcome Trust and Royal Society Sir Henry Dale Fellowship 218537/Z/19/Z.

## Author contributions

M. J. L., R. P., H. S., P. M. M., E. O-Z., T. J. A. M., K. P., A. M. O-P., C. B., J.M. J-G, R. E. D., M. F., conducted and designed experiments, acquired and analysed data and wrote the manuscript; M. H. analysed data and wrote the manuscript; G. B., R. P. G., S. P., A. W. S. and H. W. contributed to data and sample acquisition; P. C., M. T. K. I., A. P., E. M. and E. C. provided clinical samples; K. D., J. M-S. and M. S-H. contributed to design and execution of the study; M. A., E. P., H. E. M. contributed to experimental design, supervision, data interpretation and wrote the manuscript; R. B., J. E. and M.Z. contributed to execution of the study and provided samples; M. H. M. and S. N. contributed to design and supervision of the study; R. T. M-N. designed the study and experiments, contributed to data acquisition, interpretation and analysis and wrote the manuscript.

