## Supplemental Material for "Resilient SARS-CoV-2 diagnostics workflows including viral heat inactivation"

**Supplemental Methods**

**2019-nCoV detection probes and primers**

*Centers for Disease Control and Prevention (CDC) recommended probes and primers:* 2019-nCoV_N1-P (FAM-ACCCCGCATTACGTTTGGTGGACC-BHQ1), 2019-nCoV_N1-F (GACCCCAAAATCAGCGAAAT), 2019-nCoV_N1-R (TCTGGTTACTGCCAGTTGAATCTG), 2019-nCoV_N2-P (FAM-ACAATTTGCCCCCAGCGCTTCAG-BHQ1), 2019-nCoV_N2-F (TTACAAACATTGGCCGCAAA), 2019-nCoV_N2-R (GCGCGACATTCCGAAGAA), RP-P (FAM-TTCTGACCTGAAGGCTCTGCGCG-BHQ1), RP-F (AGATTTGGACCTGCGAGCG), and RP-R (GAGCGGCTGTCTCCACAAGT) were ordered from Integrated DNA Technologies (2019-nCoV CDC EUA Kit). Each primer/probe set (N1, N2, RNAseP) comes premixed at the recommended concentrations by the CDC.

*Charité/World Health Organization (WHO)/Public Health England (PHE) probes and primers:* RdRP_SARSr-P2 (6FAM -CAggTggAACCTCATCAggAgATgC- BBQ) were ordered from TIB MOLBIOL (Germany). RdRP_SARSr-F2 (GTGARATGGTCATGTGTGGCGG) and RdRP_SARSr-R1 (CARATGTTAAASACACTATTAGCATA) were ordered from Eurofins. We did not employ RdRP_SARSr-P1 (6FAM-CCAggTggWACRTCATCMggTgATgC- BBQ) except for Supplementary Figure 2 - as this probe is not specific for SARS-CoV-2 but generic for coronaviruses. Probes were HPLC-purified by the manufacturer. W is A/T; R is G/A; M is A/C.

**MagMAX RNA isolation using MagMAX™-96 Total RNA Isolation Kit (AM1830)**

After heat treatment/no heat treatment of nasopharyngeal swab within Class I MSC of CL-3 lab, 100 µl of sample was transferred to 1.5mL tubes and 300 µl TRIzol™ Reagent (Thermofisher 15596018) added. Samples were vortexed and incubated at room temperature for 5 mins. 40 µl chloroform was added, samples vortexed and incubated for a further 5 mins. Samples were transferred to a CL-2 lab for further processing and spun at 12000g at 4°C for 10 mins. 100ul of the upper aqueous phase was then transferred to a 96-well plate, 50 µl 100% isopropanol added, and samples vortexed/shaken for 1 min. RNA binding beads (Thermofisher Scientific) were first vortexed to resuspend, then 10ul was added to each sample, and the plate vortexed/shaken for a further 3 mins. The plate was placed onto a 96-well magnetic stand (Thermofisher Scientific) for approximately 2 mins, until the supernatant was completely clear. All of the supernatant was removed carefully, without disturbing the beads. 150 µl Wash 2 (Thermofisher Scientific) was added and the plate vortexed/shaken for 1 min. The plate was placed on the magnet until supernatant clear, and the supernatant removed. This step was repeated, using another 150 µl Wash 2. Once the last supernatant was removed the plate was vortexed/shaken for 2 minutes to dry the beads. 50 µl Elution Buffer (Thermofisher Scientific) was added to each sample and the plate vortexed/shaken vigorously for 3 mins. The plate was placed on the magnet, and once clear the supernatant containing total RNA was removed and transferred to a clean 96-well plate.

### Supplemental Figures and Tables

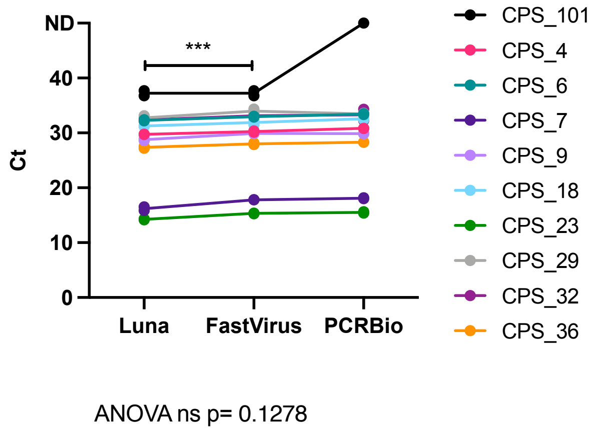


**Supplemental Figure 1. Comparison between three RT-qPCR Master Mix kits with N1 primer-probes.** RNA extraction with the QIAamp kit was done for a set of ten swab samples, previously classified as positive (CPS) by the diagnostics lab. RT-qPCR mixes were done according to each kit manufacturer's indications, maintaining the concentrations of primer-probes between the different mixes. For all panels, dots represent each individual technical duplicate, line connects the average of replicates. Appearance of one dot in samples is due to very tight qPCR replicates. Normality was assessed using D'Agostino & Pearson test prior to analysing the datasets employing ANOVA. ***: p<0.001

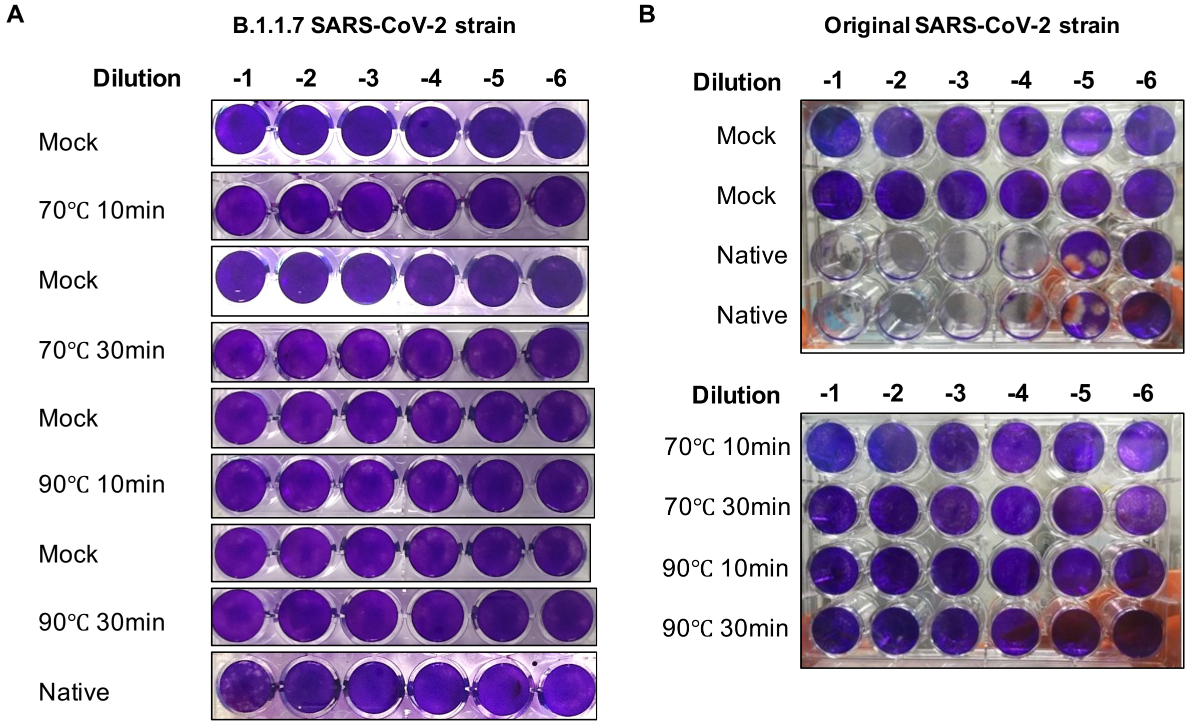


**Supplemental Figure 2. Plaque assays showing heat inactivation of the B 1.1.7 and original SARS-CoV-2 strains.** Viral stocks for B 1.1.7. **(A)** or original **(B)** strains were heat inactivated at 70°C or 90°C for 10 or 30 min, diluted and plaque assays on Vero-6 cells were performed. Note: plaques in B 1.1.7 infected cells are smaller as compared to the original strain.

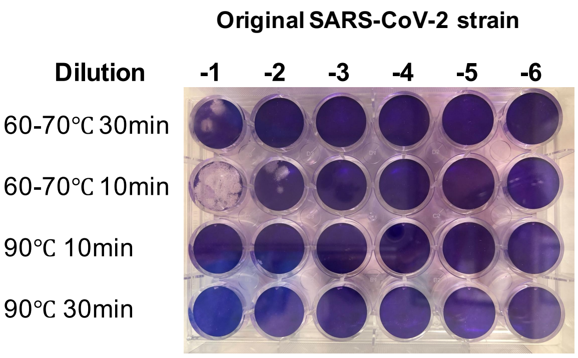


**Supplemental Figure 3. Plaque assays showing partial heat inactivation of the original SARS-CoV-2 strain.** Viral of the original SARS-CoV-2 strains were heat inactivated at 70°C or 90°C for 10 or 30 min; however, instead of using a digital thermometer we relied on the water bath temperature and later evaluated that it was set at lower than 70°C and at around 62-63°C. Plaques can be observed at the highest concentration of virus (3 x $10^5$ pfu/mL). These data show that employing less than 70°C is not safe for heat inactivation of samples, particularly of those with high viral titres. .

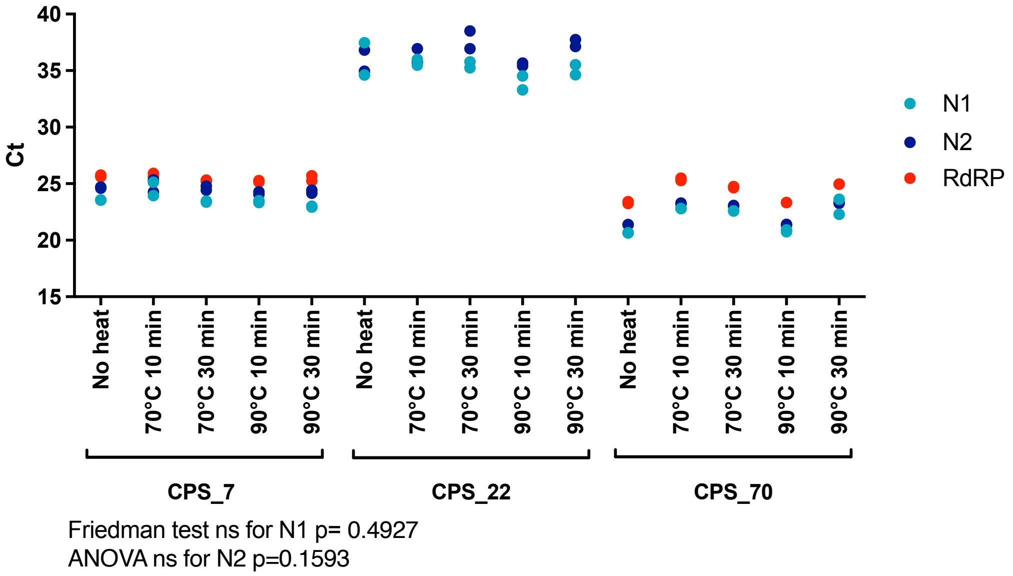


**Supplemental Figure 4. Heat inactivation of nasopharyngeal swab samples and RNA extraction using MagMax (ThermoFisher Scientific) extraction kit (A)** Six positive samples were subjected to different temperatures and incubation times as indicated. RNA was extracted using MagMax (ThermoFisher Scientific). RT-qPCR run with the three different primer-probe sets (N1, N2 and RdRP) with FastVirus Master Mix. Dots represent each individual technical duplicate.

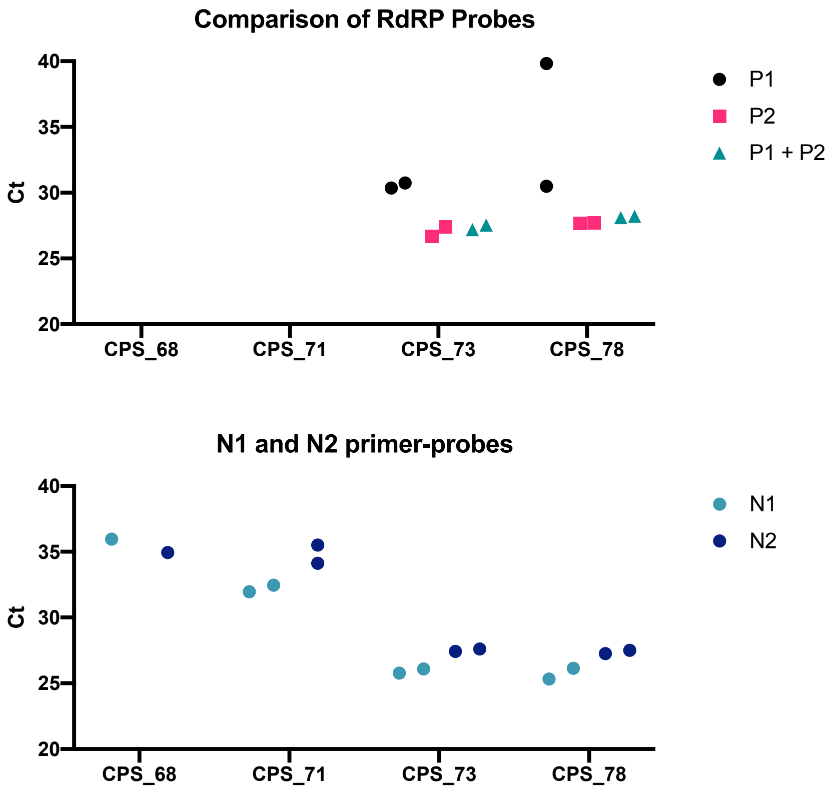


**Supplemental Figure 5: Comparison of RdRP primer-probe sets (A)** Four positive samples assessed employing the combinations of RdRP primer-probe sets (same forward and reverse, different probe combinations). RNA was extracted using QiAmp and RT-qPCR employing FastVirus Master Mix. **(B)**. Confirmation of the positivity of these samples (as per previous clinical diagnostics) employing the N1 and N2 primer-probe sets on the same samples, using Luna Master Mix. Dots represent each individual technical duplicates. For CPS_68 and N1 and N2 primer-probes we only obtained one well of amplification.

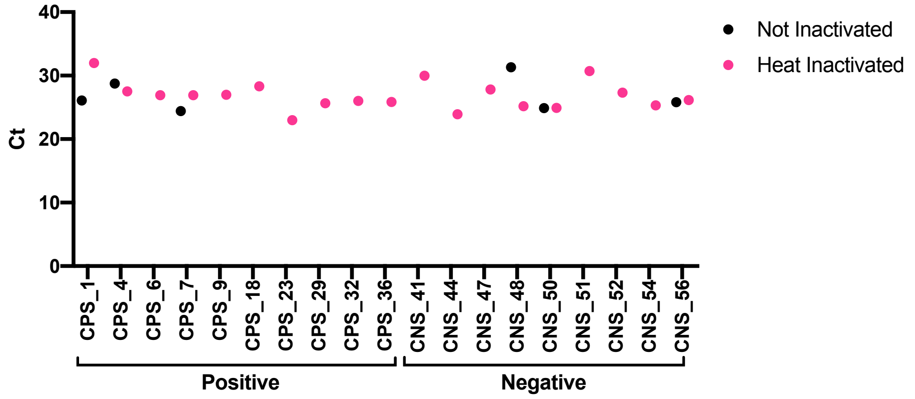


**Supplemental Figure 6. RNAseP in samples tested in Figure 5C.** Samples were tested for RNAse P for each donor except for CNS_55 where there was no RNA left to assess. All samples had detectable RNAse P. We have observed very high Ct values for RNAse P in some water controls related to bad amplification curves which were considered no amplification.

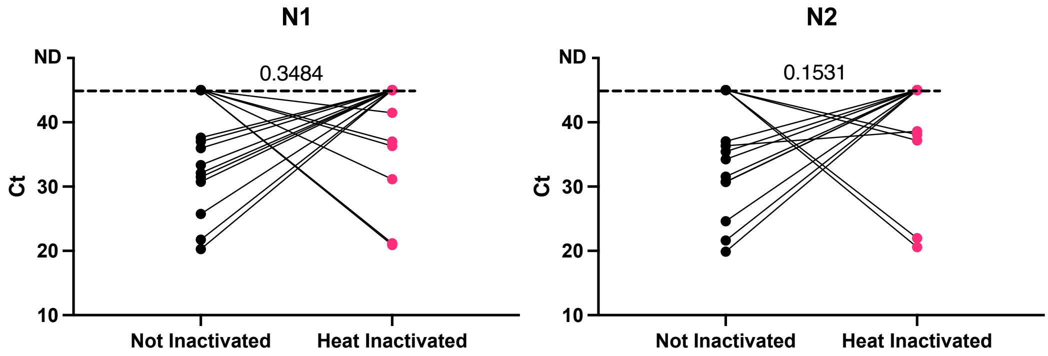


**Supplemental Figure 7. Outliers from Figure 5D.** Samples heat inactivated vs non inactivated that were detected in one or other treatment were plotted to establish if a Ct cut-off establishes detection. We were not able to determine a Ct cut-off above which samples became undetectable upon heat or vice versa, with certain samples being better detected upon heat and others (more) undetected upon heat.

| | **N1** | | **N2** | |
|---|---|---|---|---|
| | **Non-Inactivated** | **Heat Inactivated** | **Non-Inactivated** | **Heat Inactivated** |
| **Minimum** | 14.41 | 14.80 | 14.17 | 15.13 |
| **25% Percentile** | 19.82 | 20.41 | 19.82 | 20.94 |
| **Median** | 23.88 | 23.93 | 23.74 | 24.95 |
| **75% Percentile** | 30.76 | 32.89 | 30.95 | 33.23 |
| **Maximum** | 50.00 | 50.00 | 50.00 | 50.00 |
| **Range** | 35.60 | 35.21 | 35.83 | 34.88 |
| **Mean** | 26.46 | 27.76 | 26.13 | 28.06 |
| **Std. Deviation** | 9.141 | 9.998 | 8.608 | 9.577 |
| **Std. Error of Mean** | 0.9744 | 1.066 | 0.9124 | 1.015 |
| **Number of samples** | 88 | 88 | 89 | 89 |

**Supplemental Table 1**. Statistics of samples in Figure 5D. Undetermined samples were randomly assigned a value of 50. Non amplified samples in both treatments are not included.

| | **qPCRBIO Probe 1-Step Go Lo-ROX** *CDC Primers* | **TaqMan Fast Virus 1-Step** *RdRp Primers* | **TaqMan Fast Virus 1-Step** *CDC Primers* | **Luna Universal Probe** *CDC Primers* |
|---|---|---|---|---|
| **Reaction mix buffer** | 5 µL | 5 µL | 5 µL | 10 µL |
| **RT enzyme** | 2 µL RTase Go | (included in buffer) | (included in buffer) | 1 µL Luna WarmStart |
| **Primer Forward** | 1.5 µL (All premixed by IDT) | 1.2 µL (10 µM) RdRP_SARSr-F2 | 1.5 µL (All premixed by IDT) | 1.5 µL (All premixed by IDT) |
| **Primer Reverse** | | 1.6 µL (10 µM) RdRP_SARSr-R1 | | |
| **Probe** | | 0.2 µL (10 µM) RdRP_SARSr-P2 | | |
| **Water** | 5 µL | 7 µL | 8.5 µL | 2.5 µL |
| **RNA template** | 5 µL | | | |

**Supplemental Table 2**: List of components for the different one-step RT-qPCR reagents used.

| *qPCRBIO Probe 1-Step Go Lo-ROX (PCR Biosystems)* | | | |
|---|---|---|---|
| **Step** | **Temperature** | **Time** | **Cycles** |
| Reverse Transcription | 45 °C | 10 minutes | 1 |
| RT inactivation/initial denaturation | 95°C | 2 minutes | 1 |
| Denature | 95°C | 5 seconds | 50 |
| Anneal/extend | 60°C | 30 seconds | 50 |
| *TaqMan Fast Virus 1-Step Master Mix (Applied Biosystems)* | | | |
| **Step** | **Temperature** | **Time** | **Cycles** |
| Reverse Transcription | 50 °C | 5 minutes | 1 |
| RT inactivation/initial denaturation | 95°C | 20 seconds | 1 |
| Denature | 95°C | 3 seconds | 50 |
| Anneal/extend | 60°C | 30 seconds | 50 |
| *Luna Universal Probe One-Step RT-qPCR (NEB)* | | | |
| **Step** | **Temperature** | **Time** | **Cycles** |
| Reverse Transcription | 55 °C | 10 minutes | 1 |
| RT inactivation/initial denaturation | 95°C | 1 minutes | 1 |
| Denature | 95°C | 10 seconds | 50 |
| Anneal/extend | 60°C | 30 seconds | 50 |

**Supplemental Table 3**: Cycling modes for the different one-step RT-qPCR reagents used (reaction volume is always 20 µL)
